# Clinical characteristics of patients with suspected and verified PE: A single-center prospective cohort study

**DOI:** 10.1101/2025.06.04.25328957

**Authors:** Michael Marks-Hultström, Kazal Hawez, Nabil Chowdhury, Mediha Becirovic-Agic, Esin Özcelebi, Annelie Barrueta Tenhunen, Mohamad Nasir, Mikael Åberg, Christian Rylander, Miklos Lipcsey, Oskar Eriksson, Henrik Isackson, Anders Larsson, Gerhard Wikström

## Abstract

In this prospective cohort study, we included 196 patients presenting with dyspnea and suspected pulmonary embolism (PE) at the emergency department of Uppsala University Hospital, Sweden, between June 2019 and July 2022. All patients underwent CT for PE confirmation. Blood samples were collected and stored in a biobank, allowing for comprehensive biomarker analysis.

Of the 196 patients, 89 (45.4%) were diagnosed with PE. Patients with confirmed PE showed significantly elevated levels of D-dimer (median 5.7 mg/L [IQR 2.3-14) vs. 1.0 mg/L [0.4-1.8], p=1.2E-14) and CRP (41 mg/L [16–139] vs.7.2 mg/L [1.5-47], p=1.6E-7), indicating thrombotic and inflammatory activity. Cardiac biomarkers, including Troponin I and NT-pro-BNP, were also significantly higher in the PE group, reflecting cardiac strain. Interestingly, emergency room vital parameters and comorbidities were largely similar. On the other hand, there were notable differences in management, with PE patients more likely to be hospitalized and received thrombolysis more often. Patients were risk assessed using the PE Severity Index (PESI) and according to the European Society of Cardiology 2019 guidelines for PE. Patients with PE had a substantially increased mortality that remained after adjustment for comorbidities (OR: 6.0 (95% CI 2.3-15.3).

The results highlight inflammation as a central component of PE pathophysiology, which may serve as both a precipitant and a response to embolic events. Biomarkers including D-dimer, CRP, and cardiac markers enhance diagnostic accuracy and can guide management in patients with suspected PE, aligning with clinical needs in emergency care settings. Further investigation into the interaction between inflammation and coagulation in PE is important to improve risk stratification and targeted treatment approaches as mortality remains high even after diagnosis.

## Introduction

Pulmonary Embolism (PE) is a potentially life-threatening condition with high mortality that often presents with unspecific symptoms such as dyspnea and tachycardia, making rapid and accurate evaluation essential in the emergency setting[1]. Computed tomography (CT) imaging, particularly contrast-enhanced CT pulmonary angiography (CTPA), remains the gold standard for PE diagnosis[2]. Treatment is guided by the risk for early death assessed from hemodynamic parameters, right ventricular function and levels of biomarkers[2]. Despite successful initial treatment many patients go on to develop chronic thromboembolic pulmonary hypertension (CTEPH) [3] and post-thrombotic syndrome (PTS) with chronic dyspnea and functional limitations[4].

Previous studies of PE severity have highlighted several biomarkers, such as D-dimer[5], C-reactive protein (CRP)[6], and cardiac-specific injury markers like Troponin I and T[7, 8]. The level of D-dimer is known to correlate with thrombotic activity, while increased CRP levels suggest an inflammatory response, which may be either a consequence of or a contributing factor to thrombus formation[9, 10]. Elevated cardiac biomarkers such as Troponin I or T as indicators of cardiac injury and NT-pro-BNP as a marker of myocardial strain are a response to right ventricular load by PE[11]. Nevertheless, since comorbidities and patient characteristics influence these biomarkers their utility in the emergency setting warrant additional investigation.

The aim of the present study was to characterize the clinical and biomarker characteristics and to establish cut-off levels of selected biomarkers for prediction of PE among patients presenting with dyspnea and being referred for CTPA with a suspicion of PE. In addition, we wanted to investigate how these biomarkers were associated with acute management strategies, including echocardiography, thrombolysis, and hospitalization decisions, providing insight into their use in guiding treatment. Our hypothesis was that patients with confirmed PE exhibit a specific pattern of elevated biomarkers including D-dimer, CRP, and Troponin I and T, and that these correlate with assessed risk and outcome. Such findings could support a more refined, biomarker-based approach to early PE management.

## Methods

This study was conducted at the emergency department of Uppsala University Hospital in Uppsala, Sweden, and the protocol was approved by the Swedish Ethical Review Authority (initial approval Dnr: 2018/334, with a revision Dnr: 2021-00698). We included patients who were referred to CTPA on a primary suspicion of. Mortality data was accessed Jan. 24, 2025 giving a minimum of 2.5 years of follow-up. All patients provided written informed consent before inclusion and the study was performed in accordance with the Helsinki Declaration and its revisions[12].

All patients underwent a diagnostic CTPA using standard protocols for radiation exposure and iodine contrast infusion. Patient risk of early death from PE was retrospectively classified according to the PE Severity Index (PESI)[13, 14] and according to the guidelines issued by the European Society of Cardiology (ESC)[2] (Supplemental table S1). For the latterrisk classification right ventricular dysfunction was identified from the radiology report and a Troponin I above 14 ng/L or Troponin T above 40ng/L was considered elevated according to the local normal ranges. Presence of ECG features known to correspond to presence of pulmonary embolism by a blinded investigator (see Supplemental Methods).

Blood samples were collected as part of standard clinical practice upon each patient’s arrival in the emergency department. Routine chemistry included biomarkers, reflecting coagulation status, inflammation, cardiac strain, and other biochemical parameters relevant to assessing PE at the discretion of the treating physician.

### Statistics

Data are presented as n (%) for categorical parameters and mean±SD for continuous parameters, or median (IQR) for variables with skewed distributions. The study size was determined to achieve a power >80% to detect a binomial difference between the groups at a 0.05 significance level. Patients with missing data in any variable were excluded for the analysis of that variable. The risk of chance difference was calculated using Chi-square for categorical parameters, Student’s T-test for normally distributed continuous parameters and Wilcoxon Signed Rank test for skewed parameters. Kaplan-Meier survival analysis was performed to compare survival probabilities between groups, and the log-rank test was conducted to assess statistical significance between the survival distributions.

Additionally, a Cox proportional hazards model was used to evaluate the effect of PE on mortality adjusted for age, sex, and the number of comorbidities. The results are presented as hazard ratios (HR) and 95% confidence intervals (CI). Two-way ANOVA was used to distinguish the association of age with D-dimer and CRP levels. To compare the diagnostic performance of different cutoffs for plasma D-dimer levels we calculated an optimal cutoff using the Youden method[16] and compared it to the traditional population-based cutoff of 0.5 mg/L, and the age-adjusted cutoff suggested by the European Society of Cardiology (ESC) where 0.5 is used below the age of 50, and D-dimer / age above the age of 50[2]. We then compared the number of false positive and false negative results and calculated sensitivity and specificity. To investigate the added diagnostic performance of CRP we analyzed the area under the receiver operator curve (AUC ROC) for CRP alone, and for a logistic model combining D-dimer and CRP including complete cases. P < 0.05 was considered significant. Statistical analyses were performed using R version 4.2.3.

## Results

A total of 196 patients were included between June 12, 2019, and July 6, 2022. Out of these 89 (45.4%) were confirmed to have PE and 107 (54.6%) were found to have no PE. There was no loss to follow-up (Figure 1). The mean age in the PE group was at 64±16 years, compared to 61±18 years in the no-PE group. The proportion of men was 55% among patients with PE and 42% among those without PE (Table 1). No comorbidities differed significantly between groups, but while hypertension was common in both groups (36% in PE and 32% in No PE). Some less common comorbidities showed variation between groups. For example, 5 % of PE patients had a recent COVID-19 infection and 13 % of no PE patients 13%, and 23% of PE patients had a malignancy diagnosed compared to 16% of no PE patients. However, new malignancy was diagnosed in 16% of PE and 19% of no PE patients. Other comorbidities, such as cardiovascular disease, heart failure, lung disease, and history of thromboembolism showed similar distributions in both group. Further, the total number of comorbidities did not differ between the groups.

**Figure 1.**
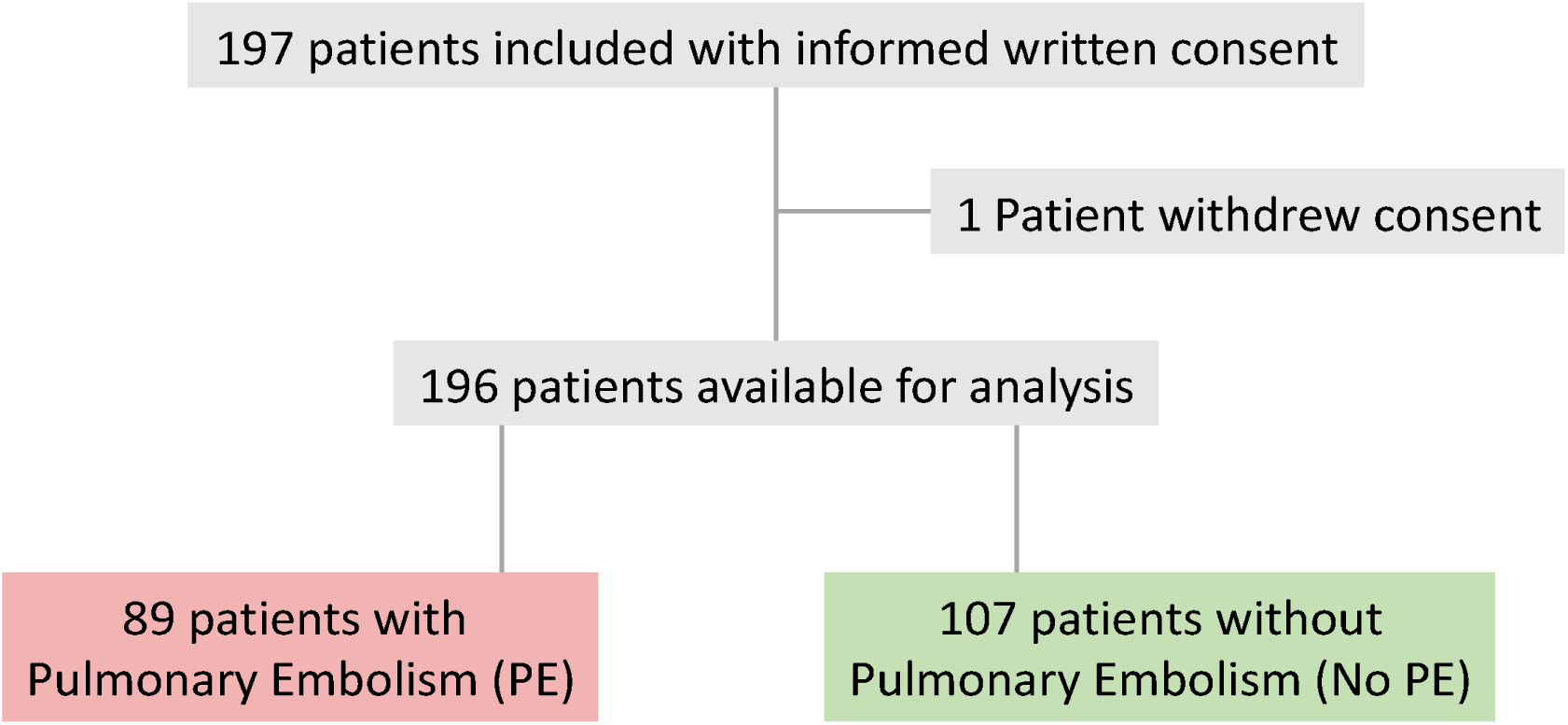

**Table 1.** Demography of 196 patients with suspected pulmonary embolism (PE) showed no major differences in age, sex or comorbidities between patients who were confirmed to have PE and those without. New malignancy refers to neoplasms identified after the diagnosis of PE, and was not included in the calculation of the number of comorbidities. Data are given as n(%) or mean (SD). The risk of chance difference was calculated using Student’s T-test for age, and Chi-square for categorical data. P < 0.05

|  | No-PE | PE | p |
| --- | --- | --- | --- |
| n | 107 | 89 |  |
| Age (mean±SD) | 61 (18) | 64 (16) | 0.35 |
| Male sex (%) | 45 (42) | 49 (55) | 0.095 |
| **Comorbidities** |  |  |  |
| Recent COVID-19 (%) | 14 (13) | 4 (4.5) | 0.068 |
| Hypertension (%) | 34 (32) | 32 (36) | 0.64 |
| Atrial fibrillation (%) | 10 (9) | 4 (4.5) | 0.30 |
| Cardiovascular disease (%) | 21 (20) | 11 (12) | 0.24 |
| Heart failure (%) | 15 (14) | 7 (7.9) | 0.26 |
| Kidney disease (%) | 3 (2.8) | 3 (3.4) | 1 |
| Lung disease (%) | 27 (25) | 17 (19) | 0.39 |
| Diabetes (%) | 17 (16) | 10 (11) | 0.46 |
| Malignancy (%) | 17 (16) | 20 (23) | 0.32 |
| New Malignancy (%) | 20 (19) | 14 (16) | 0.72 |
| Psychiatric disease (%) | 2 (1.9) | 2 (2.2) | 1 |
| Neurological disease (%) | 4 (3.7) | 1 (1.1) | 0.48 |
| Gastrointestinal disease (%) | 2 (1.9) | 3 (3.4) | 0.84 |
| Thromboembolism (%) | 13 (12) | 12 (14) | 0.95 |
| **Number of comorbidities (%)** |  |  | 0.093 |
| 0 | 24 (22) | 26 (29) |  |
| 1 | 31 (29) | 31 (35) |  |
| 2 | 22 (21) | 16 (18) |  |
| 3 | 18 (17) | 5 (6) |  |
| 4 | 10 (9) | 9 (10) |  |
| 5 | 2 (1.9) | 0 (0.0) |  |
| 6 | 0 (0.0) | 2 (2.2) |  |

The emergency room vital parameters were largely similar, indicating minimal differences in initial clinical presentation between the groups (Table 2). Oxygen saturation was slightly lower in PE patients (mean 95±4%) compared to no PE patients (97±3%; p = 0.007). The need for supplemental oxygen was also comparable, with 23% of PE patients and 18% of no-PE patients receiving oxygen therapy (p = 0.48), and the average amount of supplemental oxygen was similar between groups. Respiratory rate, an important indicator of respiratory distress, showed no difference between groups (20±5, vs. 20±6, p = 0.6). Heart rate was elevated in PE compared to control, while systolic and diastolic blood pressure were similar between groups, further suggesting that PE patients did not exhibit drastically different vital signs compared to No PE. This was further emphasized by there not being a difference of the number of patients beyond traditional severity cut-offs for systolic blood pressure and heart rate (Supplemental Table S2). ECG analysis showed most patients in sinus rhythm, and no difference between groups in the number of patients with atrial fibrillation or flutter. Right ventricular strain and clockwise rotation beyond V4 were more common in PE (Supplemental Table S3, Supplemental Figure S2), while left bundle branch block (LBBB) was more common in no PE (Supplemental Table S3).

**Table 2.** Clinical characteristics and outcomes of 196 patients with suspected pulmonary embolism (PE) showing minor but significant differences in heart rate and saturation at admission. AKI: acute kidney injury. ICU: intensive care unit.

|  | No-PE | PE | p | missing n (%) |
| --- | --- | --- | --- | --- |
| n | 107 | 89 |  |  |
| Saturation % (mean±SD) | 97±3 | 95±4 | 0.007 | 1 (0.5) |
| Supplemental oxygen n(%) | 19 (18) | 20 (22) | 0.481 | 1 (0.5) |
| Oxygen L/min (mean±SD) | 3.8±3.1 | 4.8±5.1 | 0.474 | 1 (0.5) |
| Respiratory Rate (mean±SD) | 20±6 | 20±5 | 0.609 | 3 (1.5) |
| Heart Rate (mean±SD) | 85±21 | 94±21 | 0.002 | 7 (3.6) |
| Systolic Blood Pressure (mean±SD) | 140±24 | 136±23 | 0.231 | 1 (0.5) |
| Diastolic Blood Pressure (mean±SD) | 84±16 | 83±14 | 0.686 | 3 (1.5) |
| Echocardiography n(%) | 23 (21) | 57 (64) | <0.001 | 0 |
| Thrombolysis n(%) | 0 (0) | 20 (23) | <0.001 | 0 |
| **Level of care n(%)** |  |  |  |  |
| Ward | 53 (50) | 84 (94) | <0.001 | 0 |
| ICU | 1 (1) | 0 (0) |  |  |
| Home | 51 (48) | 5 (6) |  |  |
| Missing | 2 (2) | 0 (0) |  |  |
| **AKI (n%)** |  |  | <0.001 |  |
| 0 | 42 (39) | 46 (52) |  |  |
| 1 | 5 (5) | 7 (8) |  |  |
| 2 | 1 (1) | 0 (0) |  |  |
| 3 | 0 | 0 |  |  |
| missing baseline | 59 (55) | 36 (40) | 0.14 | 95 (48) |
| **Mortality n(%)** |  |  |  |  |
| 30 days | 0 (0.0) | 6 (7) | 0.021 | 0 |
| 365 days | 6 (6) | 19 (21) | 0.002 | 0 |

The acute management of the PE group included more frequent echocardiography (64% compared to 21% in the no-PE group, p < 0.001). Thrombolysis was administered to 20 (21%) of the patients with PE. Almost all patients with PE were hospitalized (94%), whereas 50% of no-PE patients discharged to the home. Finally, mortality was substantially higher in the PE group both at 30 and 365 days after diagnosis (Table 2, Figure 2A), and remained after adjustment for age, sex and the number of comorbidities in a COX proportional hazards model (Figure 2B). Age and the number of comorbidities were significant predictors of mortality, while sex showed no significant association (Figure 2B).

**Figure 2.**
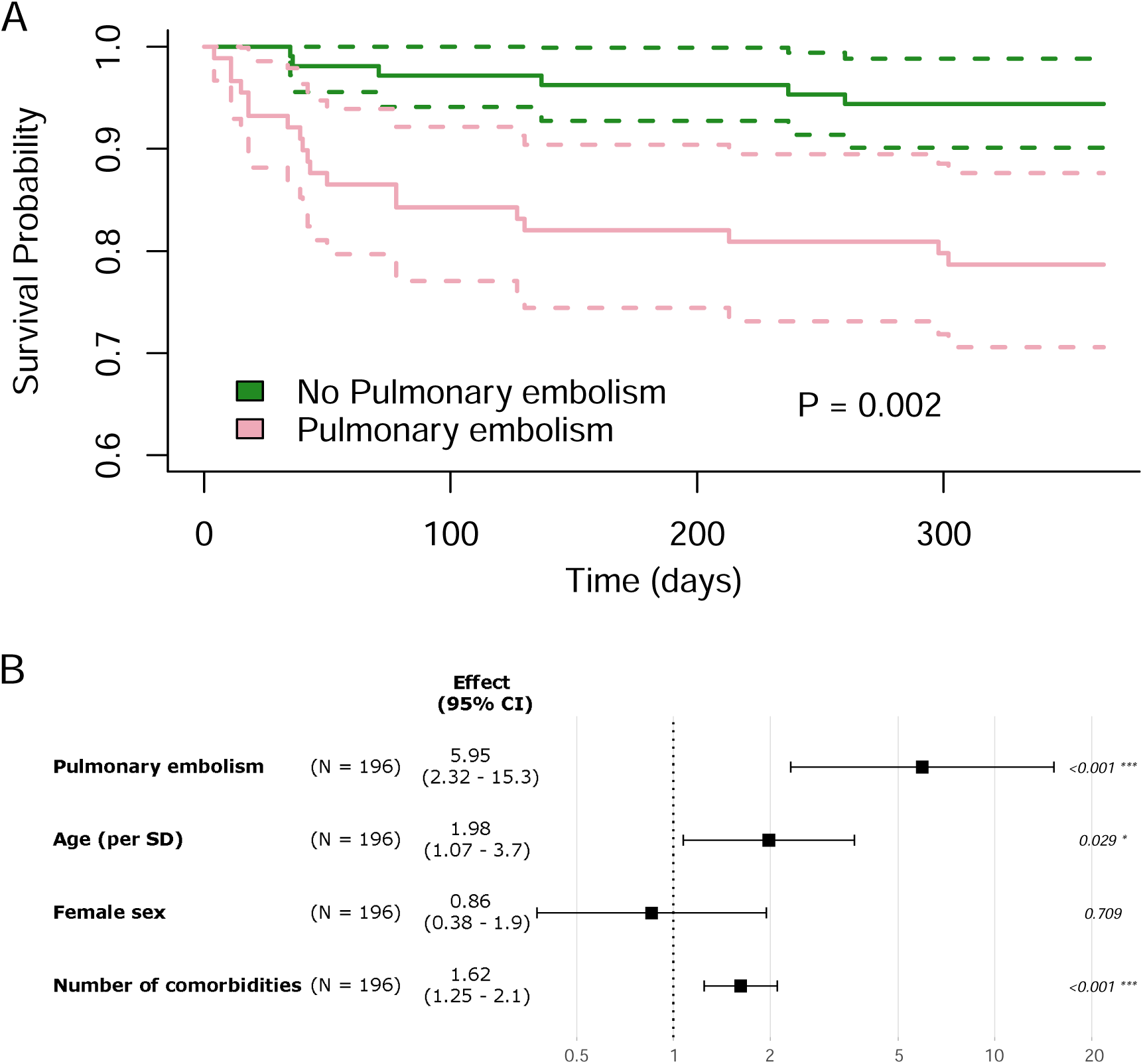
**A**: One-year survival of 196 patients with suspected PE (PE) who were diagnosed with PE (pink) or not (green). P-value is based on a Log-rank test. **B:** COX proportional hazards model for mortality within one year after PE adjusting for age (scaled to mean 0, standard deviation 1), sex and the number of comorbidities (Table 1).

Coagulation markers show significant differences between patients with confirmed PE and those without. The fibrin D-dimer levels were markedly higher in the PE group (median 5.4 [IQR 2-14]) compared to the no-PE group (median 0.9 [IQR 0.4-1.9], p=1.2E-14). On the other hand, prothrombin time and platelet count did not differ significantly between groups (Table 3). The C-reactive protein (CRP) was significantly elevated in the PE group (median 41 [IQR 16-139] vs. no-PE group 7.2 [1.5-47], p=1.6E-7). Cardiac markers exhibited clear differences between the groups. Troponin I and T, markers of myocardial injury, were both significantly elevated in the PE group (Table 3). Additionally, NT-pro-BNP levels were higher in the PE group suggesting greater cardiac strain among PE patients.

**Table 3.** Admission day biochemistry indicative of pulmonary embolism (PE), inflammation, and end-organ damage. Statistics are given as mean±SD or median [IQR]. The risk of chance difference was calculated using Student’s T-test for normally distributed variables and the Wilcoxon Signed Rank test for skewed variables. P < 0.05 was considered significant.

|  | No PE | PE | p (T-test) | p (Wilcoxon) | Missing n (%) |
| --- | --- | --- | --- | --- | --- |
| **n** | 107 | 89 |  |  |  |
| Fibrin D-Dimer (median [IQR]) | 0.9 [0.4-1.9] | 5.4 [2.0-14] |  | 1.24E-14 | 12 (6) |
| CRP (median [IQR]) | 7.2 [1.5-47] | 41 [16-139] |  | 1.62E-07 | 23 (12) |
| Prothrombin time (mean±SD) | 1.1±0.2 | 1.1±0.1 | 0.34 | | 43 (22) |
| Trombocyter (mean±SD) | 256±81 | 244±79 | 0.32 | | 22 (11) |
| Min pH (mean±SD) | 7.42±0.10 | 7.41±0.14 | 0.66 | | 134 (68) |
| Max pCO<sub>2</sub> (mean±SD) | 5.11±0.97 | 5.10±1.19 | 0.97 | | 134 (68) |
| Min pO<sub>2</sub> (mean±SD) | 9.08±3.54 | 8.13±2.69 | 0.26 | | 134 (68) |
| Max Laktat (median [IQR]) | 1.45 [1.2-2.4] | 1.1 [1.05-1.3] |  | 0.15 | 134 (68) |
| Hemoglobin (mean±SD) | 124±20 | 111±27 | 0.44 | | 22 (11) |
| Troponin I (median [IQR]) | 7.6 [2-31] | 38 [4.6-284] |  | 2.1e-3 | 85 (43) |
| Troponin T (median [IQR]) | 8 [5-16] | 42 [18-77] |  | 1.1e-4 | 146(74) |
| NT-pro-BNP (median [IQR]) | 188 [57-973] | 859 [150-3520] |  | 3.0e-3 | 2 (1) |
| Creatinine (mean±SD) | 82±30) | 85±29 | 0.47 | | 28 (14) |
| Cystatin-C (mean±SD) | 1.1±0.5 | 1.3±0.4 | 0.36 | | 170 (87) |
| ALAT (mean±SD) | 0.6±0.7 | 0.5±0.4 | 0.61 | | 97 (49) |
| ASAT (mean±SD) | 0.8±0.5 | 0.7±0.5 | 0.53 | | 105 (54) |
| Glucose (mean±SD) | 7.4±3.5 | 8.0±3.6 | 0.39 | | 46 (23) |

Renal and metabolic markers were largely similar across the groups, with no significant differences in maximum creatinine, cystatin-C, or glucose levels. Similarly, liver enzyme levels, including aspartate aminotransferase (ALAT) and alanine aminotransferase (ASAT), showed no substantial differences, suggesting that liver and kidney function were not acutely impaired by PE in this cohort. Acid-base balance parameters such as pH, pCO₂, and pO₂ also did not differ significantly (Table 3).

With regard to risk stratification, there were 18 patients in the intermediate-high and high-risk groups for PESI, and 20 intermediate-high or high risk according to the ESC guidelines (Table 5). Further, the PESI score was an predictor of mortality but not the ESC risk classification (Table 4). Thrombolysis was administered to 11 (55%) out of 20 patients in the ESC Intermediate-High Risk and High-Risk groups, and to 9 patients in the Low and Intermediate-Low Risk groups (Table 4). The cardiac specific risk score variables were different between PE and no PE patients, underscoring their importance for risk stratification and diagnosis (Supplemental table S2).

**Table 4.** Risk classification of 89 patients with pulmonary embolism (PE) according to PE Severity Index (PESI risk class) and the European Society of Cardiology (ESC) 2019 Guidelines for PE (ESC risk class), and whether they were treated with thrombolysis according to guidelines, and one-year mortality.

| **PESI risk class (%)** | **n (%)** | **Thrombolysis (%)** | **1 year mortality (%)** |
| --- | --- | --- | --- |
| I (Low) | 22 (25) | 5 (19) | 0 (0) |
| II (Low) | 25 (28) | 6 (19.4) | 1 (4) |
| III (Intermediate-Low) | 24 (27) | 5 (17.2) | 6 (25) |
| IV (Intermediate-High) | 10 (11) | 4 (28.6) | 5 (50) |
| V (High) | 8 (9) | 0 (0) | 7 (87.5) |
| **ESC risk class (%)** |  |  |  |
| Low Risk | 36 (40) | 2 (6) | 5 (13.9) |
| Intermediate-Low Risk | 33 (37) | 7 (21) | 12 (36.4) |
| Intermediate-High Risk | 19 (21) | 10 (53) | 2 (10.5) |
| High Risk (Massive PE) | 1 (1) | 1 (100) | 0 (0) |

We examined the relationship between age and fibrin D-dimer (Figure 3A) and CRP (Figure 3B) levels respectively in patients with and without confirmed PE. Two-way ANOVA demonstrated a difference in D-dimer levels between patients with PE and those without, with overall higher levels in the PE group. Furthermore, in patients without PE, there was a significant positive association between age and D-dimer levels (R² = 0.36, p = 1.96E-10), indicating that D-dimer levels increase with age even in the absence of PE. In contrast, among patients with confirmed PE, D-dimer levels did not show a significant correlation with age (R² = 0.013, p = 0.17).

**Figure 3.**
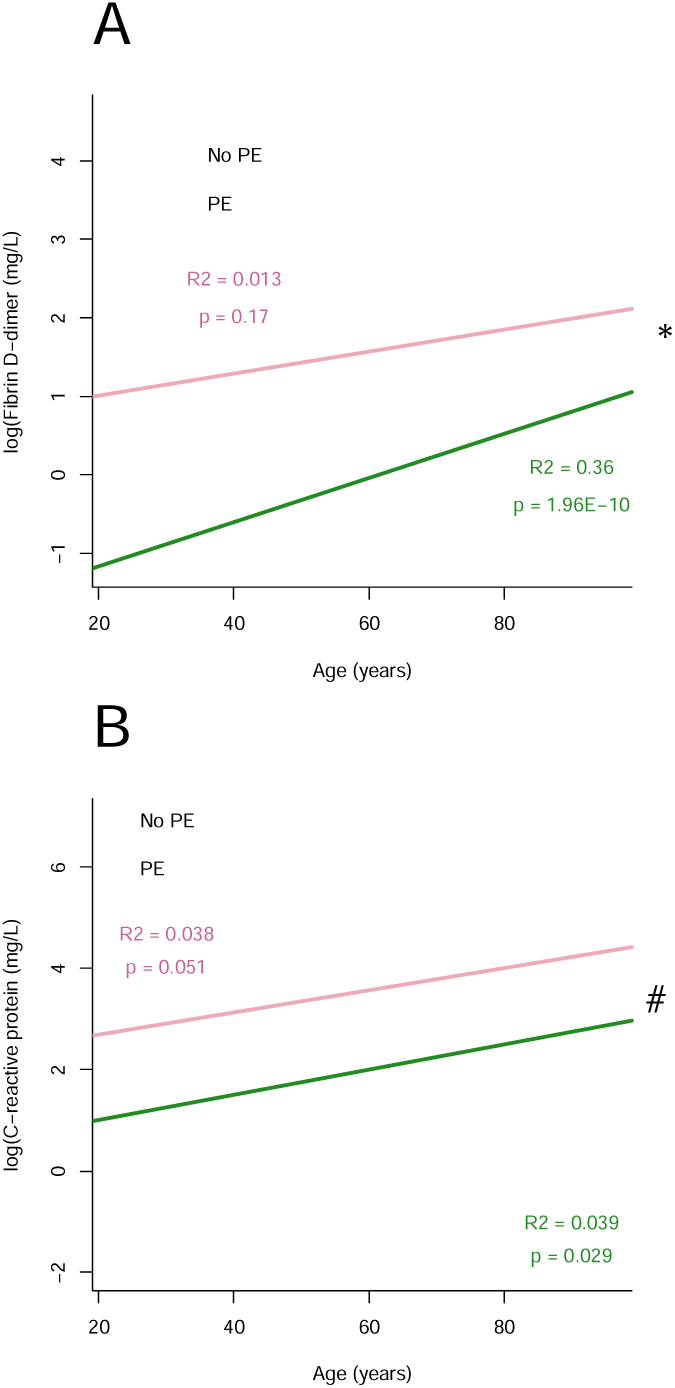
Relationship between age and log-transformed fibrin D-dimer levels (A), and CRP levels (B) in patients diagnosed with PE (pink dots) and those without PE (gray dots). Two-way ANOVA revealed a statistically significant difference in D-dimer levels between patients with PE and those without (p = 7.25E-10, indicated by *). Additionally, among patients without PE, there is a significant increase in D-dimer levels with increasing age), suggesting age-related elevation in D-dimer independent of PE. In contrast, in patients with confirmed PE, there was no significant association between D-dimer levels and age. CRP was also higher in patients with PE (p = 0.00045, indicated by #), and increased with age but not differently between patients with PE and without.

Since the median of the no-PE group was substantially higher (0.9 mg/L) than the traditional population-based cutoff (0.5 mg/L) we investigated the diagnostic performance of different cutoffs for D-dimer in this cohort. D-dimer alone has a high diagnostic performance with an AUC ROC of 86% with a Younden threshold for maximizing the AUC of 2.45 mg/L in this cohort (Figure 4A). However, this cutoff has an low sensitivity of 75%, producing 21 false negative results while still leaving 17 false positives. The traditional cutoff of 0.5 mg/L gives a sensitivity of 98% with only 2 false negatives, but proportionally worse specificity 28% with 72 false positives. An age-adjusted threshold marginally improved sensitivity to 96%, but also improved specificity to 29%.

**Figure 4.**
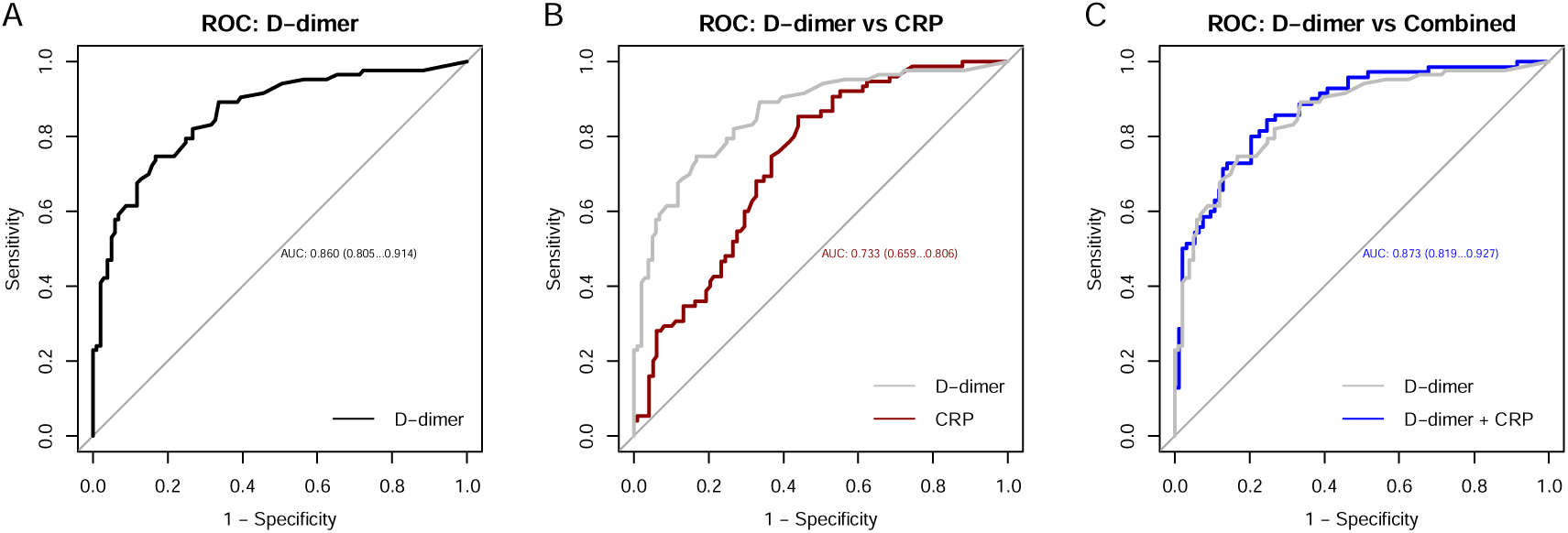
**A:** Diagnostic performance of D-dimer in 196 patients with suspected pulmonary embolism (PE) illustrated as a receiver operator curve (ROC). Area under the curve (AUC) 86%, optimal threshold according to Youden: 2.45 mg/L. **B**: ROC of CRP with D-dimer for reference, showing a lower AUC (73%), but also a markedly worse specificity, as expected. **C**: A logistic model combining D-dimer and CRP only marginally improves the AUC (87%), suggesting that CRP does not add clinically meaningful information for diagnosis of PE. ROC: receiver operator curve, AUC: area under the curve, CRP: c-reactive protein.

Finally, the added value of CRP in diagnostic performance was investigated using a logistic model. CRP alone had an AUC ROC of 73% (Figure 4B) with an optimal threshold of 9.5 mg/L. A combined logistic model of D-dimer and CRP only marginally improved the performance over D-dimer alone (AUC ROC 87% vs. 86%, Figure 4C). Troponin showed a similarly poor performance and did not add diagnostic accuracy over D-dimer alone (Supplemental figure S1).

## Discussion

The main findings of this single-center cohort study of patients with suspected PE underscore the distinct biochemical profiles of patients with confirmed PE compared to those without in an emergency department setting. While the clinical presentation both or symptoms and vital signs were similar in both groups, patients with PE presented with significantly elevated fibrin D-dimer and C-reactive protein (CRP) levels, indicative of increased thrombotic and inflammatory activity, which is consistent with the pathophysiological response to PE as reported earlier[6]. Additionally, cardiac biomarkers such as Troponin I and NT-pro-BNP were markedly higher in PE patients, suggesting greater cardiac strain due to the right ventricular pressure overload caused by the embolic obstruction as well recognized[7]. Despite treatment mortality remained high in the PE group.

The similar distribution of comorbidities in patients with and without confirmed PE suggests that while certain conditions, such as hypertension and malignancy, were prevalent among PE patients, the overall comorbidity profile did not distinctly differentiate those with PE from those without. This finding indicates that while these comorbidities are common in patients presenting to the emergency department, they are not predictive of PE in this setting, which has not been widely recognized previously. This lack of distinct comorbid profiles in PE cases underlines the complexity of identifying PE based solely on patient history and highlights the need for further investigation into other potential predictive factors specific to PE. Identifying such factors could improve risk stratification in emergency settings, aiding clinicians in prioritizing diagnostic resources and tailoring initial management.

Parameters such as oxygen saturation, respiratory rate, and blood pressure were comparable in both groups, underscoring the difficulty of diagnosing PE based on clinical signs alone. This homogeneity in vital signs illustrates the limitations of relying solely on initial physical presentation to distinguish PE from other causes of dyspnea or chest discomfort. Consequently, these findings support the critical role of additional diagnostic tools, such as specific biomarkers and imaging, in accurately identifying PE. Biomarkers like D-dimer, when used in conjunction with imaging, can help overcome the limitations of unspecific vital signs, allowing for a more definitive and timely diagnosis of PE in the emergency department.

Previous studies have shown that while ECG is a key tool in assessing suspected pulmonary embolism (PE), a normal ECG can still be found in 9–30% of cases, and findings such as sinus tachycardia, RBBB, QRS-axis deviations, clockwise rotation of the transition zone, ST-depressions, S1Q3T3 pattern, P-pulmonale, and atrial arrhythmias have all been variably reported[15]. In our study, the heart rate was higher in PE, consistent with circulatory stress. Clockwise rotation was significantly more common in PE than in no PE, aligning with earlier reports of its association with right ventricular strain. Interestingly, atrial fibrillation and flutter were more frequent in the non-PE group, though this difference disappeared when only newly diagnosed cases were considered—likely reflecting more widespread anticoagulant use in known arrhythmia patients, which may reduce PE risk. Other classic ECG findings associated with PE did not differ significantly between groups in our cohort. Notably S1Q3T3 that has been considered pathognomonic for PE occurred in both PE and no PE patients and did not show any difference between groups.

Most patients with PE in this cohort presented with lower-risk characteristics, with few patients exhibiting the hemodynamic instability or severe hypoxemia that would typically necessitate intensive care or more aggressive interventions. Consequently, the lack of intensive care admissions is consistent with a clinical picture that is not immediately life-threatening in these patients.

However, our survival analysis shows that PE was associated with a substantially higher both short-term and long-term mortality. This may be indicative of unrecognized underlying conditions or complications, especially as it was robust to adjustment for covariates.

The elevated level of C-reactive protein (CRP) in PE patients compared to controls, indicated a heightened inflammatory response among those with confirmed PE. This may represent a systemic reaction to thrombus formation in the pulmonary vasculature or, alternatively, may suggest that underlying inflammation or infection could play a role in predisposing individuals to PE. The fact that elevated CRP was not associated with other signs of infection or diagnosis of infection suggests that is can be considered a part of a broader response to vascular occlusion as has been described previously[6].

The finding of increasing D-dimer with age, particularly in patients without PE, underscores the importance of considering age when interpreting D-dimer levels in the diagnosis of PE (PE)[17]. In patients without PE, D-dimer levels showed a significant positive correlation with age, suggesting that age-related increases in D-dimer could lead to false-positive results if age-adjustment is not considered. Conversely, in patients with confirmed PE, D-dimer levels were generally elevated but did not correlate significantly with age, indicating that PE itself drives high D-dimer levels irrespective of age. The median of the no-PE group (1.0mg/L) was higher than the normal reference level of 0.5 mg/L, meaning that the laboratory’s standard reference interval classifies a large proportion of elderly, no PE patients as positive, leading to unnecessary imaging. However, we show that an age-adjusted threshold does not meaningfully improve specificity, and a statistically optimal threshold has too poor sensitivity to be used to rule out such a high-risk condition as PE.

Mortality as assessed using the risk scores was hard to estimate at 30 days, therefore we present 1- year mortality. Even if PESI was derived in PE patients it is a very general mortality score that relies heavily on age and chronic disease, it is therefore not surprising that it is an excellent predictor of mortality. Interestingly, the ESC risk score did not predict the risk of death well. Mortality was highest in the Intermediate-Low risk group, although there was only one High Risk patient in this cohort. Clinical management according to guidelines was moderately accurate, only 55% of patients indicated for thrombolysis according to the ESC guidelines received it. At the same time 13% of patients with low-risk scores were also treated with thrombolysis, indicating an area where clinical judgement and physiological derangement disagree. Whether this indicates a need to improve local adherence to guidelines or if there are aspects to clinical judgement not taken into account in the current guidelines was not part of the present analysis.

This study has several strengths like the prospective design, which allows for a comprehensive inclusion of both clinical and laboratory data on patients presenting with suspected PE (PE). The standardized use of CT ensured consistent diagnostic accuracy across the cohort and minimized diagnostic variability, allowing reliable classification of patients. The inclusion of a comprehensive array of biomarkers, collected as part of routine emergency department protocols and preserved in a biobank for later analysis, enables a detailed investigation into the biochemical profile of PE. This biobank support ensures the integrity and consistency of admission samples, allowing for standardized comparisons across cases. Additionally, the study’s focus on patients who underwent specific PE investigation aligns with real-world clinical settings, where physicians must distinguish PE from other causes of dyspnea among symptomatic patients, rather than from healthy individuals.

A limitation is that all patients included were already selected for PE investigation, thus there is no inclusion of healthy controls, which may restrict the generalizability of findings to other populations without dyspnea or similar symptoms. While this reflects the clinical setting of the study, it means that the data cannot establish the specificity of these biomarkers for PE as opposed to no-PE conditions without overlapping symptoms. The low number of early deaths precluded more detailed analysis of 30-day mortality which would have been more interesting for identifying PE-related deaths. Further, the clinical management of patients with and without PE differed, specifically in the rate of admission, and thus the number of follow-up samples, making investigation of the time-profile of biomarkers impossible. The non-protocolized follow-up also mean a relatively high number of missing for biochemistry that are at the clinician’s discretion. As a single-center study, the findings may not fully represent diverse healthcare settings or patient demographics. Despite these limitations, the study provides valuable insights into the diagnostic and management implications of biomarkers in a cohort that reflects a realistic clinical scenario in emergency medicine.

In conclusion, this study highlights the significant role of inflammation in the presentation and progression of PE (PE). Elevated levels of inflammatory markers, such as C-reactive protein (CRP), observed in patients with confirmed PE underscore inflammation’s integral association with the condition, whether as a contributing factor leading to thrombus formation or as a physiological response to the embolism itself. This inflammatory response may exacerbate the burden on the cardiovascular system, contributing to the observed elevations in cardiac biomarkers like Troponin I and NT-pro-BNP. Future studies investigating the interplay between inflammation and thrombosis in PE could further refine our understanding of this relationship, potentially guiding more targeted approaches in both prevention and acute management.

## Supporting information

Supplemental methods and results

## Acknowledgement

The authors thank the research nurses at the Department of Medicine and all the healthcare personnel at the Emergency Department at Uppsala University Hospital for their help with patient inclusion and data collection. The authors acknowledge the use of ChatGPT (Open AI) for code and text generation used in the drafting of this article. All text was edited and checked for correctness by the authors. The Uppsala University guidelines for the academic use of AI were followed in the preparation of the manuscript.

## Author contributions

Planning and design: MMH, GW, KH. Data collection: HI, GW, KH, NC, MMH, AL, MBA, OE. Statistical analysis: MMH, HI. Draft writing: MMH, GW, HI. Editing manuscript: All authors. Approving final draft: All authors.

## Data availability

All data is available from the authors on reasonable request after obtaining appropriate ethical permission and arranging data transfer agreement.

## Abbreviations

PE: pulmonary embolism
PESI: pulmonary embolism severity index
CTPA: computed tomography pulmonary angiography
PTS: post thrombotic syndrome
CRP: C-reactive protein
ALAT: aspartate aminotransferase
ASAT: alanine aminotransferase
ECG: electrocardiogram
S1Q3T3: prominent S-wave in lead 1, and a Q-wave and inverted T-wave in lead 3
AUC: area under the curve
ROC: receiver operator curve

