## Supplemental methods and results for "Clinical characteristics of patients with suspected and verified PE: A single-center prospective cohort study"

ECG analysis: Features analysed as indicative of PE were sinus tachycardia, P-pulmonale, right QRS-axis deviation, anterior ST-depression, clockwise rotation of the anterior transition zone in or beyond precordial lead V5, and right bundle branch block. The ECG recorded in closest proximity to study inclusion date was used. The ECG investigator was blinded until the stage of data analysis. P-pulmonale was defined as a pointed P-wave of more than 2.5 mV in lead II[15]. ECG with LBBB morphologies were excluded from electrical axis, anterior ST-depression, QRS-transition zone and S1Q3T3 -analyses. RBBB was assessed on morphology and included complete as well as incomplete (QRS-duration less than 120 ms).

**Supplemental table S1**

Comparison of the Pulmonary Embolism Severity Index (PESI) and risk assessment according to the 2019 European Society of Cardiology Guidelines for Pulmonary Embolism. SBP: systolic blood pressure, RVD: right ventricular dysfunction.

| **PESI parameter** | **Points** |  |
| --- | --- | --- |
| Age | 1 point per year |  |
| Male sex | 10 |  |
| Cancer | 30 |  |
| Heart failure | 10 |  |
| Chronic lung disease | 10 |  |
| Heart rate ≥ 110 bpm | 20 |  |
| SBP < 100 mmHg | 30 |  |
| Temperature < 36°C | 20 |  |
| Altered mental status | 60 |  |
| Arterial O₂ saturation < 90% | 20 |  |
| **PESI Risk Class** | **PESI Score** | **30-day Mortality** |
| I (Low) | ≤65 | 0–1.6% |
| II (Low) | 66–85 | 1.7–3.5% |
| III (Intermediate-low) | 86–105 | 3.2–7.1% |
| IV (Intermediate-high) | 106–125 | 4.0–11.4% |
| V (High) | >125 | 10.0–24.5% |
| **ESC 2019 Risk Class** | **Criteria** | **30-day Mortality** |
| Low Risk | No RVD, normal troponin, stable vitals | <1% |
| Intermediate-Low Risk | RVD OR only biomarker elevation | <5% |
| Intermediate-High Risk | RVD AND elevated troponin | 3–15% |
| High Risk | SBP < 90 mmHg OR shock | >15% |

**Supplemental Table S2:**

Risk score component variables in a cohort of 196 patients with suspected pulmonary embolism (PE) showing significant effects for cardiac enzymes, but not for physiological parameters, or even the pulmonary embolism severity index (PESI). SBP: systolic blood pressure, HR: heart rate.

| **Risk score variables** | **No PE** | **PE** | **p-value** |
| --- | --- | --- | --- |
| n | 107 | 89 |  |
| Troponin I > 14 or T > 40 (n (%)) | 30 (28) | 49 (55) | <0.001 |
| NT-proBNP (n (%)) | 32 (30) | 49 (55) | 0.001 |
| SBP < 100 mmHg (n (%)) | 3 (3) | 2 (2) | 1 |
| SBP < 90 mmHg (n (%)) | 1 (1) | 1 (1) | 1 |
| HR > 110 bpm (n (%)) | 13 (12) | 18 (20) | 0.178 |
| HR > 120 bpm (n (%)) | 8 (8) | 8 (9) | 0.902 |
| HR > 130 bpm (n (%)) | 6 (6) | 2 (2) | 0.412 |
| Saturation < 90% (n (%)) | 3 (3) | 4 (5) | 0.804 |
| Right ventricual dysfunction (n (%)) | 0 (0) | 24 (27) | <0.001 |
| PESI score (mean±SD) | 78±31 | 84±28 | 0.146 |

**Supplemental table S3**

ECG characteristics of 196 patients with suspected pulmonary embolism (PE), a total of 106 of 107 no PE patients and 83 of 86 PE had a recorded ECG, leaving 7 missing (3.6%).

|  | **No PE** | **PE** | **P-value** |
| --- | --- | --- | --- |
| n | 106 | 83 |  |
| Rhythm (%) |  |  | 0.455 |
| 2:1 atrial flutter | 2 ( 1.9) | 1 ( 1.2) |  |
| 3:1 atrial flutter | 1 ( 0.9) | 0 ( 0.0) |  |
| atrial fibrillation | 7 ( 6.6) | 2 ( 2.4) |  |
| pacemaker | 1 ( 0.9) | 0 ( 0.0) |  |
| sinus rythm | 95 (89.6) | 80 (96.4) |  |
| Ventricular frequency (mean (SD)) | 84.65 (21.12) | 94.23 (20.90) | 0.002 |
| Heart Rate for patients in Sinus Rythm (mean (SD)) | 82.68 (18.23) | 93.51 (19.14) | <0.001 |
| Atrial.fibrillation.or.flutter = 1 (%) | 10 ( 9.5) | 3 ( 3.6) | 0.195 |
| Novel.diagnosis.atrial.fibrillation.or.flutter = 1 (%) | 2 ( 1.9) | 2 ( 2.4) | 1 |
| S1Q3T3 = 1 (%) | 4 ( 4.3) | 9 (11.7) | 0.125 |
| P.pulmonale = 1 (%) | 5 ( 5.2) | 6 ( 7.5) | 0.741 |
| Right.ventricular.strain = 1 (%) | 23 (24.0) | 37 (46.2) | 0.003 |
| Right.axis.deviation = 1 (%) | 25 (27.8) | 25 (32.9) | 0.585 |
| Axis (mean (SD)) | 23.23 (37.22) | 24.59 (42.38) | 0.826 |
| Clockwise.rotation..beyond.or.equal.to.V4. = 1 (%) | 8 ( 9.1) | 22 (30.1) | 0.001 |
| Numeric.Precordial.transition.point (mean (SD)) | 3.20 (0.89) | 3.72 (1.20) | 0.002 |
| Clockwise.rotation..beyond.or.equal.to.V5. = 1 (%) | 4 (50.0) | 18 (81.8) | 0.202 |
| RBBB = 1 (%) | 9 ( 8.7) | 9 (10.8) | 0.799 |
| LBBB = 1 (%) | 9 ( 8.6) | 1 ( 1.2) | 0.056 |

**Supplemental Figure S2.**

ROC curves for D-dimer, Troponin in red, and a logistical model of D-dimer and Troponin showing no added diagnostic accuracy.

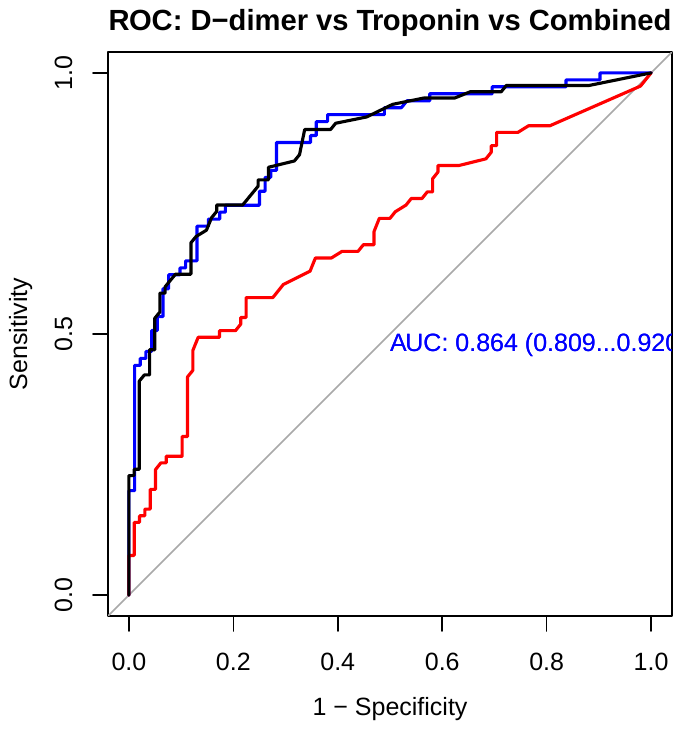

**Supplemental figure S2**

Electrical axis and precordial transition of 196 patients with supected pulmonary embolism (PE). A: Electrical axis showing no significant difference between PE and no PE patients. B: precordial transition showing a clear rotation where values beyond or equal to V4 is more common in PE.

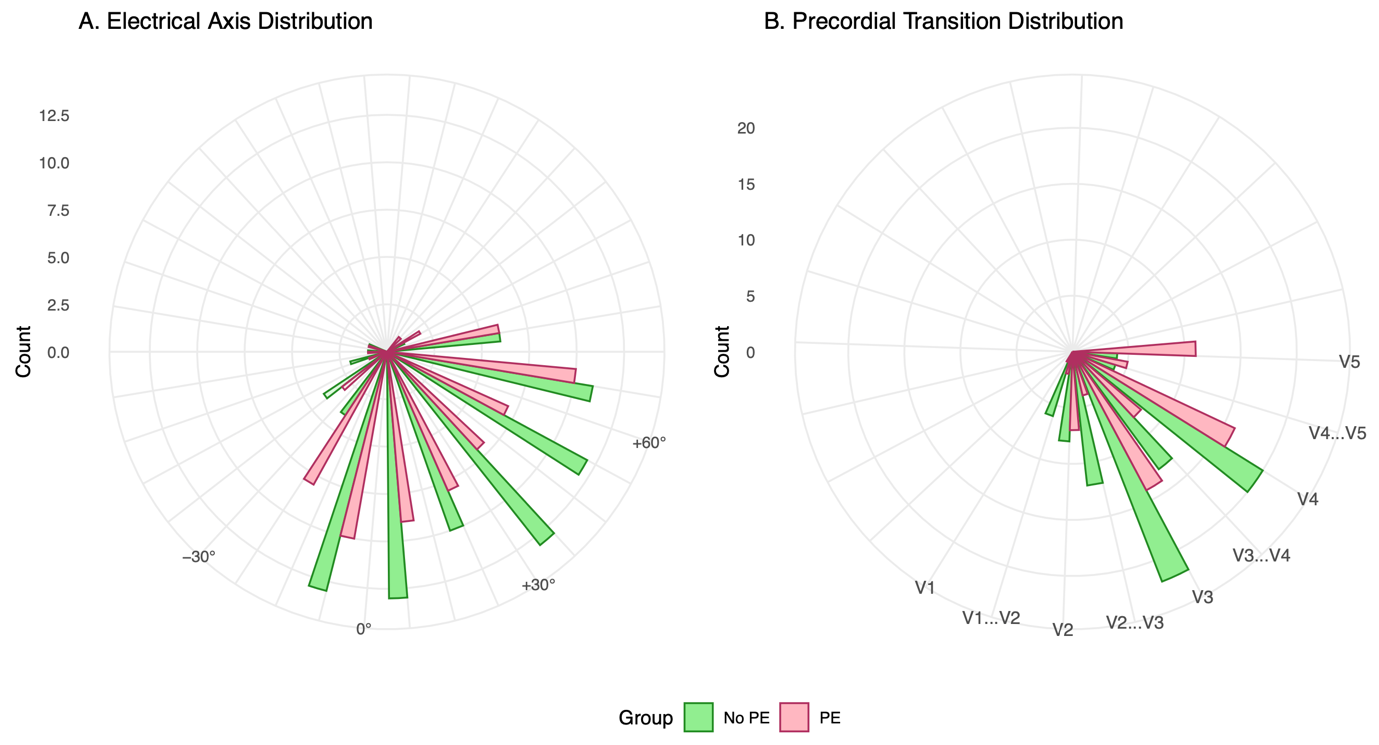
